# Neurocognition and its association with adverse childhood experiences and familial risk of mental illness

**DOI:** 10.1101/2021.11.28.21266887

**Authors:** Sai Priya Lakkireddy, Srinivas Balachander, Pavithra Dayal, Mahashweta Bhattacharya, Mino Susan Joseph, Pramod Kumar, Anand Jose Kannampuzha, Sreenivasulu Mallappagari, Suvarna Shruthi, Alen Alexander Chandy, Muthu Kumaran, Sweta Sheth, Vinutha Ramesh, Joan C Puzhakkal, S Sowmya Selvaraj, Dhruva Ithal, Vanteemar S Sreeraj, Jayant Mahadevan, Bharath Holla, Ganesan Venkatasubramanian, John P. John, Pratima Murthy, Vivek Benegal, YC Janardhan Reddy, Sanjeev Jain, ADBS Consortium, Biju Viswanath

**Affiliations:** Accelerator program for Discovery in Brain disorders using Stem cells (ADBS), Department of Psychiatry, National Institute of Mental Health and Neurosciences (NIMHANS), Bangalore, Karnataka, India

**Author notes:** **Correspondence:** Dr Biju Viswanath, Associate Professor, Department of Psychiatry, National Institute of Mental Health & Neuro Sciences (NIMHANS), Bangalore, Karnataka, India 560029; Dr Srinivas Balachander, Assistant Professor, Department of Psychiatry, ADBS, NIMHANS, Bangalore. **Funding Statement:** This research is funded by the Accelerator program for Discovery in Brain disorders using Stem cells (ADBS) (jointly funded by the Department of Biotechnology, Government of India, and the Pratiksha trust; Grant BT/PR17316/MED/31/326/2015). BV is funded by the Intermediate (Clinical and Public Health) Fellowship (IA/CPHI/20/1/505266) of the DBT/Wellcome Trust India Alliance.

**Keywords:** Childhood trauma, ACEs, neurocognition, vulnerability, familial risk, childhood deprivation, cognitive performance, childhood neglect

## Abstract

**Background:** Neurocognitive deficits are considered an endophenotype for several psychiatric disorders, typically studied in unaffected first-degree relatives (FDRs). Environmental factors such as adverse childhood experiences (ACEs) may also affect neurocognition. This study examines the effect of ACEs on neurocognitive performance in FDRs of patients with severe mental illness in order to determine whether familial risk has a moderating effect on the relationship between ACEs and neurocognition.

**Methods:** The sample consists of a total of 512 individuals composed of unaffected FDRs from multiplex families with severe mental illnesses (schizophrenia, bipolar disorder, obsessive-compulsive disorder or alcohol use disorder) and healthy controls (with no familial risk). Neurocognitive tests included processing speed (Color Trails), new learning (Auditory Verbal Learning Test), working memory (N-Back), and Theory of Mind (SOCRATIS). ACEs were measured using the WHO ACE-International Questionnaire (ACE-IQ). Regression models adjusted for age, gender and education were done to predict each neurocognitive domain by the effect of familial risk, ACE-IQ Total Score and the interaction (familial risk x ACE-IQ Total score).

**Results:** When all FDRs were examined as a group, the main effect of familial risk predicted poor performance in all domains of neurocognition (p <0.01), and the ACEs x familial risk interaction had a significant negative association with global neurocognition, processing speed & working memory. This interaction effect was driven predominantly by the familial risk of AUD. In FDRs of schizophrenia & bipolar disorder, only the main effects of familial risk were significant (working memory, theory of mind & global neurocognition), with no impact of ACEs or its interaction in both these sub-groups.

**Conclusions:** The impact of childhood adversity on neurocognition is moderated by familial risk of psychiatric disorders. Genetic or familial vulnerability may play a greater role in disorders such as schizophrenia and bipolar disorder, while the interaction between ACEs and family history may be more relevant in the case of disorders with greater environmental risk, such as substance use.

## 1. Introduction

Adverse childhood experiences (ACEs) have been studied extensively over the last decade (Struck et al., 2021), and linked to a number of negative outcomes concerning mental & physical health (Chang et al., 2019; Sonu et al., 2019). Exposure to ACEs is associated with an increased risk, and higher severity of several psychiatric disorders, including mood & anxiety disorders (Selous et al., 2020), psychosis (Carbone et al., 2019), substance use disorders (SUD) (Hughes et al., 2019) and obsessive-compulsive disorder (OCD) (Visser et al., 2014). The effects of ACEs are also understood to be transdiagnostic; we demonstrated in a previous report that even in individuals with high familial loading/genetic vulnerability, ACEs accelerated the age at onset in many of the above-mentioned disorders (Someshwar et al., 2020).

Several mechanisms are postulated to explain how ACEs might lead to these effects. Earlier research focused on the psychological & social aspects of early life stress, and how they affect attachment patterns, bonding and thus impact emotional processing (Pollak, 2008). However, it is increasingly recognized that the mechanisms that underlie the impact of ACEs are also biological. There is substantial evidence that ACEs lead to premature biological ageing, demonstrated at the cellular level by reduced telomere length (Bürgin et al., 2019; Warner et al., 2020), physiological effects such as earlier pubertal timing (Henrichs et al., 2014; Holdsworth & Appleton, 2020), and various brain changes such as smaller age-related volumes of hippocampus in individuals who report ACEs (Bick & Nelson, 2016; Dannlowski et al., 2012; Herzog et al., 2020; Vythilingam et al., 2002).

Emerging literature suggests that there may be a link between ACEs and neurocognitive functioning. The largest study to date, which analyzed data from the UK National Household Longitudinal Study (n=27,985 adults), found that early-life adversity was associated with poorer performance on word recall, verbal fluency and numerical ability tasks (Bridger & Daly, 2019). Many other studies on smaller samples have also demonstrated that adults who have experienced ACEs perform poorer in various neurocognitive domains. (Cowell et al., 2015; Hawkins et al., 2021; Kavanaugh et al., 2017; Majer et al., 2010).

However, there are also studies that suggest the opposite relationship. One such study, drawing data from the Irish Longitudinal Study on Ageing, (n=6,912 adults aged >50 years) found, surprisingly, that childhood sexual abuse (CSA) was associated with better global cognition, memory, executive function, and processing speed, despite poorer psychological health (Feeney et al., 2013). Two other studies also reported similar findings, of better working memory in adults with a history of adverse/unpredictable childhoods (Mittal et al., 2015; Young et al., 2018). Another large population-based study, the English Longitudinal Study on Ageing (n=5223), did not find any association between ACEs & memory decline (immediate and delayed recall) (O’Shea et al., 2021). A systematic review of studies looking at the association between ACEs and neurocognition in healthy individuals found a high degree of heterogeneity in methods & their findings (Su et al., 2019).

Neurocognitive deficits are considered a core feature of many psychiatric disorders. Patients with psychiatric disorders, who also have a history of ACEs have greater deficits compared to those without history of ACEs (R.-Mercier et al., 2018). A meta-analysis of 23 studies with a total of 3315 patients with psychotic disorders (Vargas et al., 2019), examined the relationship between ACEs and overall neurocognitive functioning for four subdomains (working memory, executive function, verbal/visual memory, and attention/processing speed). Only small negative associations (effect sizes between 0.05 - 0.1) were found between overall cognition, or any of the sub-domains and childhood trauma in individuals with psychotic disorders. Only a few such studies have been done in samples with other psychiatric disorders, which also suggest poor neurocognitive functioning with a history of ACEs, in alcohol use disorder (AUD) (Cheng et al., 2020) and in major depressive disorder (Dannehl et al., 2017)

In the context of psychiatric disorders, neurocognitive deficits have also traditionally been conceptualized as candidate endophenotypic markers. Endophenotypes, by definition, are quantifiable measures postulated to be stable (found preceding the onset of the disorder as well as in remission) and heritable. Research on neurocognitive functioning in clinically healthy first-degree relatives (FDRs) of probands with various psychiatric disorders have found poorer performance on various domains in comparison to controls, with considerable overlap in the deficits identified across disorders (Abramovitch et al., 2021; East-Richard et al., 2020). Neurocognitive deficits are thus postulated to represent genetic risk for psychiatric disorders, which are polygenic and have complex heterogeneous distal phenotypes. In addition to shared genetic factors, FDRs also may share environmental factors, particularly ACEs with probands. Moreover, there is fairly robust evidence that ACEs may worsen neurocognition in those with a psychiatric diagnosis, but the relationship has shown mixed results in healthy individuals. Hence, it may be important to examine how ACEs may influence neurocognitive functioning in FDRs as well.

In this study, we aimed to describe the relationship between ACEs and neurocognitive functioning in at-risk FDRs from multiplex families with major psychiatric disorders - schizophrenia (SCZ), bipolar disorder (BD), alcohol use disorders (AUD) & obsessive-compulsive disorder (OCD). A trans-diagnostic approach could help us to understand the differential effect of ACEs on the neurocognitive profiles of such syndromes. In addition, evaluating individuals from multiplex families with a pre-existing genetic loading for psychiatric disorders might inform us about the nature of neurocognitive endophenotypes and the role of ACEs in increasing neurocognitive deficits in vulnerable individuals. We hypothesized that ACEs would affect neurocognition differentially in FDRs vs healthy controls.

## 2. Methodology

### 2.1 Sample Recruitment

We used data from the Accelerator program for Discovery in Brain disorders using Stem cells (ADBS) project, an ongoing longitudinal study (Viswanath et al., 2018). The project was reviewed and approved by the Institutional Ethics Review Board and written informed consent was obtained from all individuals who were recruited.

The ADBS project includes families in whom multiple members (at least two affected FDRs in a nuclear family) are diagnosed to have a major psychiatric disorder (schizophrenia, BD, OCD, Alzheimer’s dementia, and SUD). Patients who report such a family history during their clinical evaluation are invited for participation. Familial loading is verified by applying the Family Interview for Genetic Studies (FIGS) (Maxwell, 1996), along with a pedigree using information by interviewing at least three members from the family. The psychiatric diagnosis (or lack thereof in unaffected FDRs) is corroborated by two trained psychiatrists using the Mini International Neuropsychiatric Interview (MINI) (Sheehan et al., 1998). For details about various clinical characteristics in the sample such as diagnoses, comorbidity, intra-familial co-occurrence of different syndromes, we refer the reader to our profile paper (Sreeraj et al., 2021). The healthy control group consisted of volunteers aged between 16 to 60 years who were screened for the absence of any current/lifetime psychiatric diagnosis in self as well as in any FDR as per the MINI and the FIGS.

### 2.2 Assessments

Individuals for whom complete sociodemographic, clinical information (including age of onset of the syndrome, gender, maximum education attained and psychiatric diagnosis), Adverse Childhood Experiences – International Questionnaire (ACE-IQ) form (World Health Organization, 2018) & neurocognitive test scores were available were included. Socio-economic status was assessed using the Modified Kuppuswamy Scale, which is validated in India. (Ananthan, 2021) A total of 512 participants (unaffected ‘at-risk’ FDRs & healthy controls) met these criteria from the data available in the first wave of the ADBS project.

#### 2.2.1 Adverse Childhood Experiences

ACEs were assessed by trained post-graduate clinical psychologists, psychiatric social workers, or psychiatrists, all of whom underwent regular training in the use of the instrument. The WHO ACE-IQ questionnaire (World Health Organization, 2018), consists of 31 questions across 13 subdomains: physical abuse; emotional abuse; contact sexual abuse; alcohol and/or drug abuser in the household; incarcerated household member; household member with a psychiatric condition or suicidality; household member treated violently; one or no parents, parental separation or divorce; emotional neglect; physical neglect; bullying; community violence; and collective violence.

A total ACE exposure score was calculated by adding the total number of subdomains where any adversity was reported irrespective of the frequency at which it might have occurred (aka “Binary Score”). A total ACE severity score was also calculated by adding the total number of subdomains where adversity was reported to have occurred above a predefined frequency threshold (aka “Frequency Score”). This threshold differed by the subdomain of ACE reported (e.g. contact sexual abuse only requires being touched sexually once, but emotional abuse requires being screamed at many times). The total ACE exposure score is sensitive and identifies the occurrence of any adversity. The total ACE severity score is specific and only detects more severe and potentially impactful adversity. Therefore, the total ACE severity score was used for this analysis. Details regarding the prevalence & severity of ACEs in our sample have been reported previously. (Someshwar et al., 2020)

#### 2.2.2 Neurocognitive Tests

All neurocognitive tests were administered, in a quiet room free of distraction, during a single session that lasted approximately 40 to 50 minutes on the forenoon of the assessment day. Trained clinical psychologists with a M.Phil degree in clinical psychology administered all the tests. The following tests were administered:

1. **Color Trails (CT) Tests** (D’Elia et al., 1996) A and B, were used as measures of processing speed, sustained attention and cognitive flexibility. Scores were obtained for total time taken to complete the tests (in seconds) and number of errors in misses and wrong selection. Combining both, a measure of “effective time-per-circle” was calculated using the formula [TT/CC+TE/TC], where TT is the Total time taken, CC is number of circles completed, TE is Total Errors committed, and TC is total circles in the test (ie., 25 for CT-A and 50 for CT-B).
2. The **Auditory N-back** (1-back & 2-back) tests were used to measure verbal working memory (Kirchner, 1958). Thirty randomly ordered consonants common in Indian languages were presented (S. L. Rao et al., 2004). Scores for the total number of omissions and commission were recorded. The main outcome measure was accuracy, which was calculated using the formula [1 – (number of commission errors + number of omission errors)/total trials] x 100.
3. **Rey’s Auditory Verbal Learning Test (RAVLT)** was used to measure verbal learning and memory (Schmidt, 1996). Two lists A and B with 15 different words from familiar objects were presented over five trials. Scores were obtained for the total numbers of words correctly recalled over all the five trials (learning score) and the number of words recalled correctly in the recall trial.
4. **Social Cognition Rating Tools in Indian Setting (SOCRATIS)** was used to assess social cognition in the Indian socio-cultural setting (Mehta et al., 2011). Subtests assessing second order theory of mind (ToM) from SOCRATIS were used, since some previous studies (Sayin et al., 2010) have shown that even though basic ToM abilities of OCD patients are generally preserved, they might show reduction in their “advanced” or “second order” ToM abilities. Adapted versions of two false belief stories (Ice-cream man task (Perner & Wimmer, 1985) and hidden banana task (Stone et al., 1998) and two Irony detection stories (Drury et al., 1998) were administered from which a second order theory of mind index (Mehta et al., 2011) was calculated as the proportion of correct responses.

### 2.3 Statistical analysis

Descriptives of the sociodemographic details and ACE scores in the sample were done using mean/standard deviations and n/percentage. As the FDRs groups were not mutually exclusive, i.e, an individual FDR could be “at-risk” for multiple diagnoses, no statistical analyses were done to compare the FDRs groups between each other. Normality was checked for all the variables by examining histograms and Q-Q plots. Variables that were found to have a non-normal distribution were transformed using automated algorithms in the “bestNormalize” package in R (Peterson, 2021).

To reduce type I error, we performed a confirmatory factor analysis to reduce 10 neurocognitive variables into 4 latent domains - processing speed, working memory, new learning/immediate recall and theory of mind. These in-turn were reduced into a “global neurocognitive ability” latent factor within the same model. The details about the model specification and its fit indices are shown in Supplemental Figure 1 & Supplemental Table 1. The latent factor scores were then extracted for each subject, and these were used as the outcome (dependent) variables for all subsequent analyses.

As an initial step, we performed a separate linear regression to look at the effect of ACEs in the whole sample, without accounting for familial risk, to predict each neurocognitive domain. The planned covariates were age, gender, years of education (as a proxy for IQ) and socio-economic status. In all the regression models (i.e for every neurocognitive domain), we found high multicollinearity, using the Tolerance & Variance Inflation Factor values (>3.0), and this was found to be due to high negative correlation between ACEs score and the SES score. As the main predictor of interest in our study was ACEs, SES was dropped as a covariate from all analyses.

For the main analysis, we used an interaction term (ACEs * Familial Risk), along with their main effects as the predictors of interest. The same covariates (Age, Gender & Years of Education). Separate models for each neurocognitive domain were then tested in sub-samples of FDRs for each disorder, while the controls (i.e, those having no familial risk, n=188) were constant.

We kept a p-value threshold of 0.05/5 = 0.01, as there were 5 outcome variables. All analysis was performed in R version 4.1.1 (R Core Team, 2021), using the base packages. Confirmatory factor analysis was done using the lavaan package (Rosseel, 2012).

## 3. Results

Table 1 shows the demographic details of the sample included in this analysis. As stated previously, FDRs groups were not mutually exclusive, i.e, an individual FDR could be “at-risk” for multiple diagnoses by having family members. Hence, no statistical comparisons were made between these groups. Noticeable differences are in education and socio-economic status - these were higher in controls and least in the AUD FDRs group. Similarly, the prevalence & extent of reported ACEs in the sample are depicted in Table 2. Overall, the group of FDRs of AUD report higher ACEs in nearly every domain. Also, the controls (having no FDRs with any psychiatric disorder in the family) were found to have lesser cumulative ACEs, in terms of dimensional, frequency & binary scores. However, reported sexual abuse was higher in them in comparison to all the other groups.

**Table 1.** Demographic characteristics of the sample (Total n = 512 Unaffected Individuals in the ADBS Cohort)

|  | **No FDR(s) with severe mental illnesses** | **Having FDR(s) with Schizophrenia** | **Having FDR(s) with OCD** | **Having FDR(s) with Bipolar disorder** | **Having FDR(s) with Alcohol use disorder** |
| --- | --- | --- | --- | --- | --- |
| **n** | **188** | **123** | **73** | **119** | **169** |
| Age (mean (SD)) | 37.26 (15.40) | 39.58 (14.64) | 40.23 (14.43) | 37.61 (15.88) | 37.71 (15.05) |
| Years of Education (mean (SD)) | 14.41 (8.58) | 10.50 (4.74) | 12.04 (4.55) | 11.15 (4.14) | 10.06 (4.32) |
| Gender = Male (%) | 101 (53.7%) | 63 (51.2%) | 38 (52.1%) | 65 (54.6%) | 70 (41.4%) |
| Occupation (%) |  |  |  |  |  |
| Employed | 102 (54.3%) | 77 (62.6%) | 39 (53.4%) | 72 (60.5%) | 94 (55.6%) |
| Homemaker | 21 (11.2%) | 23 (18.7%) | 19 (26.0%) | 19 (16.0%) | 37 (21.9%) |
| Retired | 13 ( 6.9%) | 3 ( 2.4%) | 3 ( 4.1%) | 4 ( 3.4%) | 3 ( 1.8%) |
| Student | 45 (23.9%) | 15 (12.2%) | 9 (12.3%) | 18 (15.1%) | 29 (17.2%) |
| Unemployed | 4 ( 2.1%) | 4 ( 3.3%) | 3 ( 4.1%) | 6 ( 5.0%) | 5 ( 3.0%) |
| Socio economic class (Kuppuswamy) (%) |  |  |  |  |  |
| Lower | 1 ( 0.5%) | 4 ( 3.3%) | 0 ( 0.0%) | 3 ( 2.6%) | 5 ( 3.0%) |
| Upper Lower | 29 (15.5%) | 38 (31.7%) | 14 (19.2%) | 36 (31.3%) | 79 (47.9%) |
| Lower Middle | 41 (21.9%) | 33 (27.5%) | 17 (23.3%) | 44 (38.3%) | 42 (25.5%) |
| Upper Middle | 71 (38.0%) | 29 (24.2%) | 26 (35.6%) | 25 (21.7%) | 32 (19.4%) |
| Upper | 45 (24.1%) | 16 (13.3%) | 16 (21.9%) | 7 ( 6.1%) | 7 ( 4.2%) |
| Kuppuswamy Score (mean (SD)) | 18.69 (7.09) | 14.43 (7.48) | 17.74 (7.34) | 13.49 (6.24) | 12.17 (5.97) |
| Marital Status (%) |  |  |  |  |  |
| Divorced or separated | 3 ( 1.6%) | 3 ( 2.4%) | 1 ( 1.4%) | 1 ( 0.8%) | 3 ( 1.8%) |
| Married | 110 (58.5%) | 80 (65.0%) | 49 (67.1%) | 71 (59.7%) | 109 (64.5%) |
| Single | 71 (37.8%) | 36 (29.3%) | 20 (27.4%) | 42 (35.3%) | 46 (27.2%) |
| Widowed | 3 ( 1.6%) | 4 ( 3.3%) | 2 ( 2.7%) | 5 ( 4.2%) | 10 ( 5.9%) |

**Table 2.** Characteristics (prevalence and severity) of the Adverse Childhood Experiences within the sample (N=512)

|  | **Controls**<br>(No FDR(s)<br>with severe<br>mental<br>illnesses) | **Having FDR(s)**<br>**with**<br>**Schizophrenia** | **Having FDR(s)**<br>**with OCD** | **Having FDR(s)**<br>**with Bipolar**<br>**disorder** | **Having FDR(s)**<br>**with Alcohol use**<br>**disorder** |
| --- | --- | --- | --- | --- | --- |
| **n** | **188** | **123** | **73** | **119** | **169** |
| **Adverse Childhood Experiences Total & Sub-domain scores (Mean(SD))** |  |  |  |  |  |
| Total (Binary) | 2.01 (1.87) | 3.17 (2.27) | 2.63 (2.12) | 3.12 (1.99) | 3.89 (2.43) |
| Total (Frequency) | 0.81 (1.18) | 2.04 (2.05) | 1.53 (1.64) | 1.88 (1.53) | 2.63 (2.19) |
| Total Dimensional Score | 6.20 (5.77) | 9.37 (8.48) | 7.84 (6.55) | 8.70 (6.71) | 12.23 (9.66) |
| Abuse Score | 1.44 (2.41) | 1.93 (3.26) | 1.81 (2.93) | 1.95 (2.72) | 2.54 (3.86) |
| Neglect Score | 2.22 (2.13) | 2.99 (3.09) | 2.86 (2.79) | 2.55 (2.79) | 4.13 (3.47) |
| Household Challenges<br>Score | 1.19 (1.86) | 3.20 (2.91) | 2.18 (2.59) | 3.21 (2.76) | 3.89 (3.31) |
| Community Challenges<br>Score | 1.35 (2.08) | 1.25 (1.86) | 0.99 (1.44) | 0.98 (1.69) | 1.66 (2.31) |
| **Types of ACEs reported (%) - Binary Score** |  |  |  |  |  |
| Physical Abuse | 45 (23.9%) | 33 (26.8%) | 20 (27.4%) | 32 (26.9%) | 58 (34.3%) |
| Emotional Abuse | 54 (28.7%) | 51 (41.5%) | 28 (38.4%) | 49 (41.2%) | 72 (42.6%) |
| Sexual Abuse | 19 (10.1%) | 3 ( 2.4%) | 2 ( 2.7%) | 0 ( 0.0%) | 5 ( 3.0%) |
| Family members with<br>substance use | 9 ( 4.8%) | 38 (30.9%) | 12 (16.4%) | 37 (31.1%) | 109 (64.5%) |
| Incarcerated household<br>member | 3 ( 1.6%) | 4 ( 3.3%) | 0 ( 0.0%) | 4 ( 3.4%) | 5 ( 3.0%) |
| Family members with<br>mental health issues | 5 ( 2.7%) | 53 (43.1%) | 26 (35.6%) | 60 (50.4%) | 40 (23.7%) |
| Domestic violence | 69 (36.7%) | 69 (56.1%) | 30 (41.1%) | 70 (58.8%) | 100 (59.2%) |
| Parents dead/divorced | 18 ( 9.6%) | 31 (25.2%) | 13 (17.8%) | 27 (22.7%) | 48 (28.4%) |
| Emotional neglect | 9 ( 4.8%) | 4 ( 3.3%) | 2 ( 2.7%) | 5 ( 4.2%) | 8 ( 4.7%) |
| Physical neglect | 34 (18.1%) | 40 (32.5%) | 22 (30.1%) | 34 (28.6%) | 100 (59.2%) |
| Bullied by peers | 26 (13.8%) | 17 (13.8%) | 14 (19.2%) | 15 (12.6%) | 28 (16.6%) |
| Community Violence | 78 (41.5%) | 42 (34.1%) | 20 (27.4%) | 36 (30.3%) | 74 (43.8%) |
| Collective Violence | 9 ( 4.8%) | 5 ( 4.1%) | 3 ( 4.1%) | 2 ( 1.7%) | 10 ( 5.9%) |
| **Types of ACEs reported (%) - Frequency Score** |  |  |  |  |  |
| Physical Abuse | 5 ( 2.7%) | 7 ( 5.7%) | 7 ( 9.6%) | 5 ( 4.2%) | 17 (10.1%) |
| Emotional Abuse | 7 ( 3.7%) | 14 (11.4%) | 7 ( 9.6%) | 10 ( 8.4%) | 22 (13.0%) |
| Sexual Abuse | 19 (10.2%) | 3 ( 2.5%) | 2 ( 2.8%) | 0 ( 0.0%) | 5 ( 3.0%) |
| Incarcerated household member | 3 ( 1.6%) | 4 ( 3.3%) | 0 ( 0.0%) | 4 ( 3.4%) | 5 ( 3.0%) |
| Domestic violence | 24 (12.8%) | 37 (30.1%) | 16 (21.9%) | 41 (34.5%) | 72 (42.6%) |
| Parents dead/divorced | 18 ( 9.8%) | 31 (25.6%) | 13 (17.8%) | 27 (23.1%) | 48 (28.6%) |
| Emotional neglect | 30 (16.0%) | 21 (17.1%) | 11 (15.1%) | 12 (10.1%) | 33 (19.5%) |
| Physical neglect | 11 ( 5.9%) | 21 (17.1%) | 8 (11.0%) | 16 (13.4%) | 62 (36.7%) |
| Bullied by peers | 5 ( 2.7%) | 5 ( 4.1%) | 2 ( 2.7%) | 3 ( 2.5%) | 2 ( 1.2%) |
| Community Violence | 8 ( 4.3%) | 12 ( 9.8%) | 5 ( 6.8%) | 7 ( 5.9%) | 20 (11.8%) |
| Collective Violence | 9 ( 4.8%) | 5 ( 4.1%) | 3 ( 4.2%) | 2 ( 1.7%) | 10 ( 6.0%) |

The results of the multiple linear regression model examining the effect of ACEs on the total sample, along with the effects of covariates are shown in Table 3. Significant effects of ACEs were found for the global neurocognition factor, and working memory (p<0.001 in both). There was a trend towards significance for processing speed (p=0.023), and new learning/recall (p=0.024). The covariates (age & gender especially) were found to be significant in predicting most domains.

**Table 3.** Impact of ACEs on the total sample (N=512) with effects of covariates (Age, Gender & Education)

| Predictor | Processing speed |  | New Learning |  | Working memory |  | Theory of mind |  | Global Neurocog factor |  |
| --- | --- | --- | --- | --- | --- | --- | --- | --- | --- | --- |
|  | B (SE) | p value | B (SE) | p value | B (SE) | p value | B (SE) | p value | B (SE) | p value |
| Age | -0.03<br>(0.002) | **<0.001** | -0.027<br>(0.002) | **<0.001** | -0.02<br>(0.001) | **<0.001** | -0.002<br>(0.001) | **<0.001** | -0.024<br>(0.001) | **<0.001** |
| Gender (Male) | 0.222<br>(0.058) | **<0.001** | -0.143<br>(0.059) | **0.015** | 0.079<br>(0.036) | **0.03** | 0.002<br>(0.009) | 0.815 | 0.097<br>(0.044) | **0.029** |
| Years of Education | 0.001<br>(0.001) | 0.077 | 0.001<br>(0.001) | 0.073 | 0.001<br>(0.001) | **0.013** | 0.001<br>(0.001) | 0.092 | 0.001<br>(0.001) | **0.014** |
| Adverse childhood experiences - Total Score | -0.069<br>(0.03) | **0.024** | -0.069<br>(0.03) | **0.023** | -0.05<br>(0.019) | **0.008** | -0.008<br>(0.005) | 0.082 | -0.062<br>(0.023) | **0.007** |

Table 4 shows the results of our main analysis, with the main effects of ACEs, familial risk & their interaction. When all “at risk” FDRs were examined as a group, the main effect of familial risk was significant for predicting poor performance in all domains (p <0.01), and their interaction had a significant negative association with global neurocognition, processing speed & working memory. The main effect of ACEs alone was not found significant for any of the neurocognitive domains. When looking at sub-groups of FDRs with risk of different syndromes, we found that main effects of familial risk of schizophrenia & bipolar disorder were found to predict poorer performance in working memory, theory of mind & global neurocognition, and there was no significant effect of the interaction for any domain of neurocognition. The interaction term was found significant only in the subgroup of FDRs at risk for AUD. for global neurocognition, processing speed and working memory. Neither the main effect of familial OCD risk, nor its interaction with ACEs was found to significantly predict any neurocognitive domain. Figure 1 shows the plots depicting the relation between ACEs score and global neurocognitive ability in the whole FDR sample (“any risk”) and sub-samples for each disorder, in comparison to controls with no risk.

**Table 4.** Results of regression analysis predicting each neurocognitive domain by the ACEs, familial risk & their interaction, after adjusting for age, gender & years of education.

| Familial Risk | Domain | Main effect of Adverse childhood experiences |  | Main effect of familial risk |  | Interaction ACEs x Familial risk |  |
| --- | --- | --- | --- | --- | --- | --- | --- |
|  |  | B (SE) | P value | B (SE) | P value | B (SE) | P value |
| **Any severe mental illness**<br>**N = 324** | Processing speed | 0.092 (0.053) | 0.079 | -0.274 (0.061) | **<0.001** | -0.19 (0.063) | **0.003** |
|  | New Learning | -0.008 (0.054) | 0.877 | -0.147 (0.062) | **0.019** | -0.056 (0.065) | 0.386 |
|  | Working Memory | 0.041 (0.032) | 0.212 | -0.186 (0.038) | **<0.001** | -0.1 (0.039) | **0.01** |
|  | Theory of Mind | 0.003 (0.009) | 0.73 | -0.029 (0.01) | **0.004** | -0.01 (0.01) | 0.323 |
|  | Global Neurocognitive ability | 0.051 (0.04) | 0.204 | -0.227 (0.046) | **<0.001** | -0.125 (0.048) | **0.009** |
| **Schizophrenia**<br>**N = 123** | Processing speed | 0.069 (0.05) | 0.169 | -0.137 (0.074) | 0.065 | -0.115 (0.074) | 0.121 |
|  | New Learning | -0.02 (0.054) | 0.708 | -0.109 (0.08) | 0.173 | 0.011 (0.08) | 0.893 |
|  | Working Memory | 0.027 (0.031) | 0.379 | -0.126 (0.045) | **0.006** | -0.038 (0.045) | 0.403 |
|  | Theory of Mind | 0.002 (0.009) | 0.788 | -0.032 (0.013) | **0.016** | -0.006 (0.013) | 0.655 |
|  | Global Neurocognitive ability | 0.034 (0.038) | 0.369 | -0.151 (0.056) | **0.007** | -0.05 (0.056) | 0.371 |
| **Alcohol Use Disorder**<br>**N = 169** | Processing speed | 0.088 (0.053) | 0.096 | -0.378 (0.072) | **<0.001** | -0.225 (0.071) | **0.002** |
|  | New Learning | -0.009 (0.053) | 0.866 | -0.212 (0.073) | **0.004** | -0.079 (0.072) | 0.274 |
|  | Working Memory | 0.04 (0.032) | 0.211 | -0.262 (0.044) | **<0.001** | -0.12 (0.043) | **0.006** |
|  | Theory of Mind | 0.005 (0.008) | 0.562 | -0.043 (0.011) | **<0.001** | -0.012 (0.011) | 0.275 |
|  | Global Neurocognitive ability | 0.05 (0.039) | 0.205 | -0.319 (0.054) | **<0.001** | -0.149 (0.053) | **0.005** |
| **OCD**<br>**N = 73** | Processing speed | 0.078 (0.045) | 0.085 | -0.087 (0.077) | 0.261 | -0.105 (0.081) | 0.194 |
|  | New Learning | -0.022 (0.053) | 0.67 | 0.123 (0.09) | 0.171 | 0.038 (0.095) | 0.689 |
|  | Working Memory | 0.029 (0.028) | 0.303 | 0.005 (0.048) | 0.921 | -0.045 (0.051) | 0.377 |
|  | Theory of Mind | 0.003 (0.008) | 0.73 | 0.009 (0.013) | 0.503 | -0.001 (0.014) | 0.93 |
|  | Global Neurocognitive ability | 0.037 (0.034) | 0.287 | 0.003 (0.059) | 0.964 | -0.055 (0.062) | 0.374 |
| **Bipolar Disorder**<br>**N = 119** | Processing speed | 0.072 (0.047) | 0.131 | -0.109 (0.07) | 0.119 | -0.139 (0.073) | 0.058 |
|  | New Learning | -0.032 (0.053) | 0.553 | -0.072 (0.079) | 0.363 | -0.009 (0.082) | 0.909 |
|  | Working Memory | 0.025 (0.03) | 0.405 | -0.107 (0.044) | **0.016** | -0.057 (0.046) | 0.22 |
|  | Theory of Mind | 0.001 (0.008) | 0.924 | -0.036 (0.012) | **0.004** | -0.017 (0.013) | 0.196 |
|  | Global Neurocognitive ability | 0.031 (0.037) | 0.392 | -0.127 (0.054) | **0.019** | -0.073 (0.057) | 0.196 |

**Figure 1.**
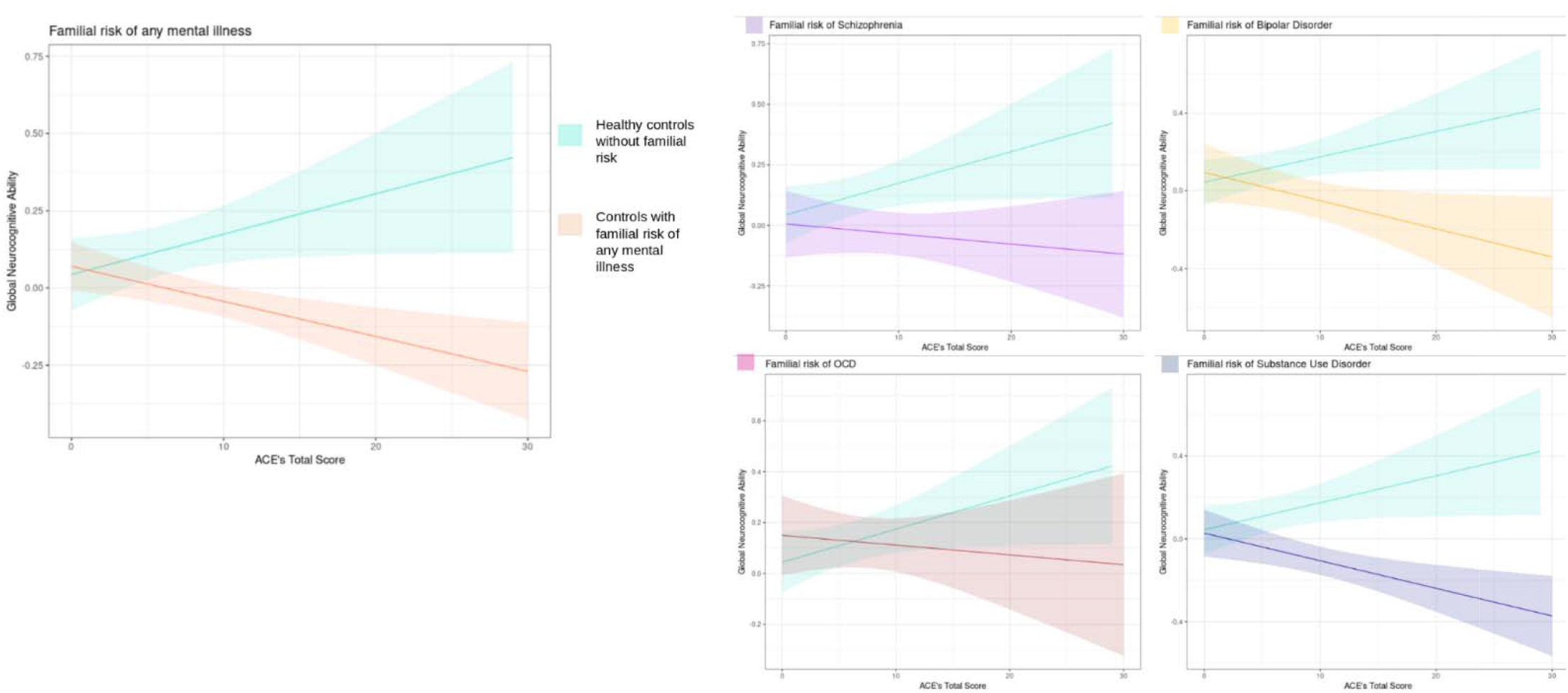
Relationship between adverse childhood experiences and global neurocognitive ability in ‘At Risk’ vs Healthy subjects.

## Discussion

The key aim of this study was to dissect the influence of childhood adversities versus genetic vulnerabilities for major mental illnesses, and how they might interact and lead to neurocognitive deficits. Neurocognitive deficits have been extensively researched from the “endophenotype” standpoint, and only recent studies have begun to look at how environmental factors might influence them. Previous such studies have looked at patients affected with schizophrenia, some with large sample sizes, but have yielded mixed results (Kasznia et al., 2021; Mansueto et al., 2018; McCabe et al., 2012; Schalinski et al., 2018; Vargas et al., 2019; Velikonja et al., 2019; Wells et al., 2020). There is hardly any research that has examined this relationship (between ACEs and neurocognition) in other disorders.

To our knowledge, we present the first study to examine how ACEs impact neurocognitive deficits in a large sample of unaffected FDRs. This approach enables us to test our hypotheses (endophenotype vs environmental), without the confounding effect of state-related factors, especially medications. Also, the FDRs included in this study were from a genetically “enriched” transdiagnostic sample of multiplex families, and were evaluated comprehensively for their family history, ACEs, as well as neurocognition. Hence, we were presumably well-poised to pick up signals due to familial risk, as well as childhood adversity across the 4 major psychiatric disorders.

The main findings of our study are as follows: When familial risk is not taken into consideration, ACEs seem to have a negative impact on all domains of neurocognition. However, including a familial risk variable into the model nullifies the effect of ACEs alone, but the interaction between the two remains significant. The domains that remained significant after multiple comparison testing were global neurocognitive ability, working memory, and processing speed. Further, this interaction effect appears to be driven predominantly by the familial risk of AUD. On the other hand, the main effects familial risk of schizophrenia and bipolar disorder were significant, with no impact of ACEs or its interaction in both these sub-groups.

The long-term effects of ACEs are undoubtedly complex, and there are likely to be myriad factors that may confer vulnerability or resilience to individuals who experience ACEs. As shown in our study, studying the effects of ACEs alone without including other moderators can be problematic. Though we found initially that ACEs have an overall negative impact on neurocognition, the effect appears to be nuanced, with differences in those who are “at risk” FDRs versus those without a family history. At the most basic level, this points out to familial risk being an important vulnerability factor for succumbing to the effects of ACEs.

The interaction term (familial risk x ACEs) was found to have a negative impact on two neurocognitive sub-domains, namely working memory and processing speed. In comparison to the other neurocognitive domains, both these have been shown to decline significantly with age (Brockmole & Logie, 2013;

McNab et al., 2015; Pliatsikas et al., 2019), as opposed to other domains such as theory of mind. This is in line with findings reported in the Longitudinal Ageing Study Amsterdam (Korten et al., 2014), where processing speed was found to decline faster over 10-year follow-up of those with ACEs & depressive symptoms, but not ACEs alone. Also, an increased rate of decline was not seen for other cognitive domains which were delayed recall and the total MMSE scores. This fits in well with the “accelerated biological ageing” hypothesis of ACEs’ impact, adding the element of specific vulnerability factors that may moderate this effect. We use familial risk as a proxy for genetic vulnerability, and one might argue that “familial risk” itself carries with it shared psychosocial and environmental factors that would confound our interpretation. Hence, it would be crucial to follow-up these studies using genotyping data, by carrying out gene x environment (G x E) analyses, or by polygenic risk scores (PRS) of various disorders instead of familial risk. Such studies have been done to predict disease risk, but not for neurocognition.

For the two severe mental disorders - schizophrenia and bipolar disorder, only familial risk was found significantly associated with poorer neurocognition, particularly in the domains of working memory and theory of mind. Neurocognitive deficits, particularly working memory as well as social cognition, are considered a core feature of psychotic disorders, and have been very consistently replicated as endophenotypes as well. Bearing in mind the high degree of genetic loading that they carry, it is possible that the effect of ACEs is “overpowered”, and is thus not found significant even as an interaction effect. A similar conclusion was drawn in the previous meta-analysis that examined correlations between ACEs and neurocognition in patients with psychosis (Vargas et al., 2019). This was attributed to factors such as medication use & disease severity, whereas in our study we find that a similar phenomenon is seen even in their FDRs. This further highlights the relevance of neurocognition as an endophenotype, especially in severe mental illnesses.

Based on twin and adoption studies, the influence of environmental & social factors is estimated to be higher in AUD, with lower heritability (Verhulst et al., 2015). It is thus possible that we found more significant signals for ACEs/interaction for the AUD risk. Also, the AUD FDRs were found to have lower education, poorer socio-economic status and may have other shared environment factors as well, which could not have been separately accounted for in our analysis, but are simply a part of “familial risk”. Hence, the endophenotype nature of the main effect of familial risk in this group needs to be interpreted with caution, as most of them could simply be better explained by other environmental factors.

The findings of our study need to be interpreted with the following limitations. Firstly, though we had a large sample of unaffected FDRs, it may be insufficient for certain individual disorders (OCD having the lowest number n=73). Also due to sample size constraints, we did not look for the effects of specific types of ACEs (e.g. abuse vs neglect). As the FDR sample was from multiplex families, many subjects had familial risk for multiple disorders, and there would be “cross contamination” across the various “familial risk” for each disorder. Again, due to sample size constraints we could not use the different familial risks & interaction with ACEs in a single model, as this would require too many main effects and interactions, resulting in difficulties in interpretation and possible Type II errors. Thirdly, our control sample (those without familial risk) were better educated, were from a higher socio-economic status than FDR groups, biases may have thus crept in even though these factors were adjusted for.

## 4. Conclusions and future directions

Our study brings forth important insights about how the impact of childhood adversity on neurocognition may be moderated by familial risk of psychiatric disorders. It also highlights how genetic/familial vulnerability may play a greater role in disorders such as schizophrenia and bipolar disorder, while the interaction may be more relevant in the case of disorders with greater environmental risk, such as substance use. Replacing the “familial risk” variable in our study with genetic markers or polygenic risk scores would help in examining specific genetic factors. Furthermore, replacing our outcome (neurocognitive performance) with a host of other markers such as personality/temperament, structural & functional imaging, or even cellular phenotypes may yield greater insights about the effects of ACEs. This will help improve how we identify and understand vulnerability & resilience factors, and perhaps better target our interventions towards them.

## Data Availability

Requests for the data related to this study may be addressed to the corresponding author(s).

## Acknowledgments

Nil

## Supplement

**Figure 1.**
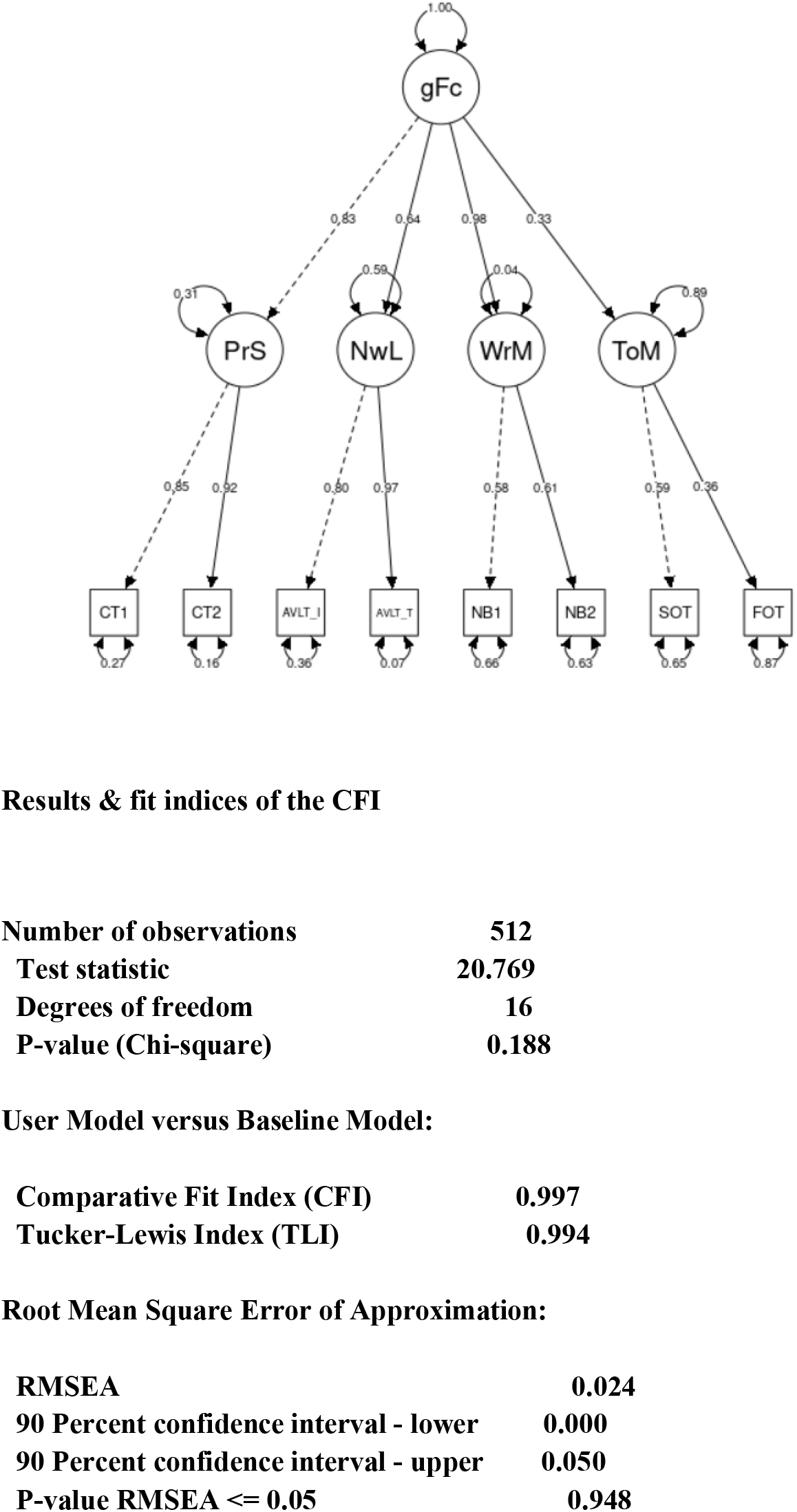
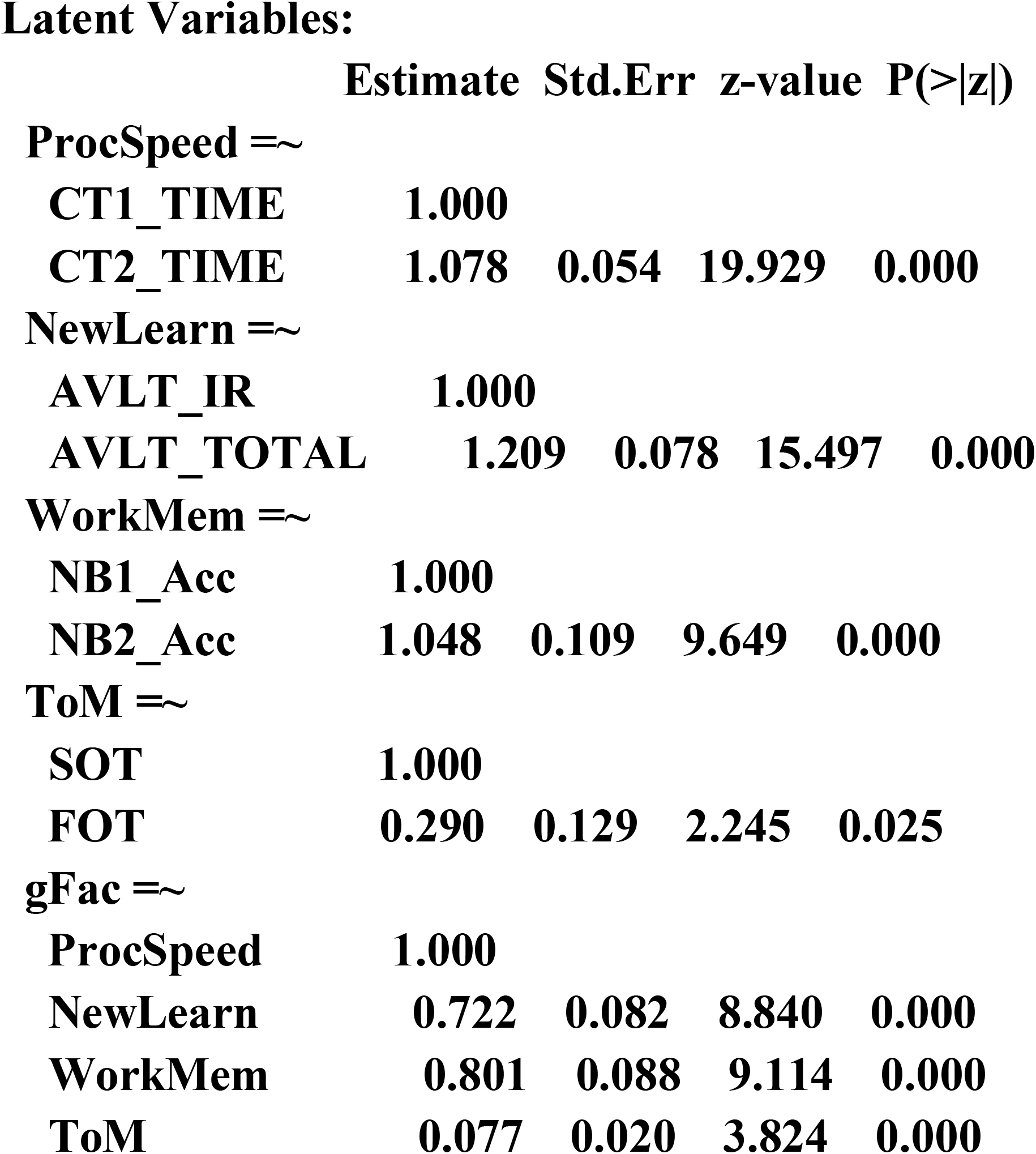
Results of Confirmatory Factor analysis of the neurocognitive variables.

